# Population-scale testing can suppress the spread of COVID-19

**DOI:** 10.1101/2020.04.27.20078329

**Authors:** Jussi Taipale, Paul Romer, Sten Linnarsson

**Affiliations:** Department of Medical Biochemistry and Biophysics, Karolinska Institutet, Stockholm, Sweden.; Applied Tumor Genomics Research Program, Faculty of Medicine, University of Helsinki, Finland and Department of Biochemistry, University of Cambridge, Cambridge, United Kingdom.; New York University

## Abstract

We propose an additional intervention that would contribute to the control of the COVID-19 pandemic, offer more protection for people working in essential jobs, and help guide an eventual reopening of society. The intervention is based on: (1) testing every individual (2) repeatedly, and (3) isolation of infected individuals. We show here that at a sufficient rate of testing and isolation, the R_0_ of SARS-CoV-2 would be reduced well below 1.0, and the epidemic would collapse. The approach does not rely on strong and/or unrealistic assumptions about test accuracy, compliance to isolation, population structure or epidemiological parameters, and its success can be monitored in real time by measuring the change of the test positivity rate over time. In addition to the rate of compliance and false negatives, the required rate of testing is dependent on the design of the testing regime, with concurrent testing outperforming random sampling of individuals. Provided that results are reported rapidly, the test frequency required to suppress an epidemic is linear with respect to R_0_, to the infectious period, and to the fraction of susceptible individuals. Importantly, the testing regime would be effective at any level of prevalence, and additive to other interventions such as contact tracing and social distancing. It would also be robust to failure, as even in the case where the testing rate would be insufficient to collapse the epidemic, it would still reduce the number of infected individuals in the population, improving both public health and economic conditions. A mass-produced, disposable antigen or RNA test that could be used at home would be ideal, due to the optimal performance of concurrent tests that return immediate results.

---

The ongoing pandemic spread of SARS-CoV-2 is the cause of widespread and accelerating outbreaks of the respiratory syndrome COVID-19. As of May 17, 2020, a total of 4 525 497 persons have been confirmed to be infected, and 307 395 have died^1^. Currently, the virus is present on all continents, spreading rapidly in Europe and the USA, and is a major threat to world order. It now seems likely that unemployment in most countries will exceed the levels reached during the depth of the Great Depression of the 1930s. Detailed models of the epidemic dynamics of SARS-CoV-2 are available that take into account population structure, and can provide estimates of the magnitude and duration of the peak of infection^2^. However, very broadly speaking, an outbreak of a novel virus in a naïve population can have only two outcomes. As long as the reproduction number R_0_ remains greater than 1, the virus spreads rapidly until most people have been infected (**Fig. 1A**), creating a temporary surge of infected individuals. If, using pharmaceutical or social interventions, R_0_ can be reduced below 1, then the epidemic collapses (**Fig. 1B**), and most people remain uninfected (but still susceptible). Because of the exponential nature of epidemics, the outcomes are nearly binary. Even when R_0_ exceeds one by only a small amount the disease spreads at an accelerating pace, whereas as soon as R_0_ falls just below one it rapidly collapses. These two outcomes correspond to two distinct strategies for epidemic control, suppression and mitigation.

**Fig. 1.**
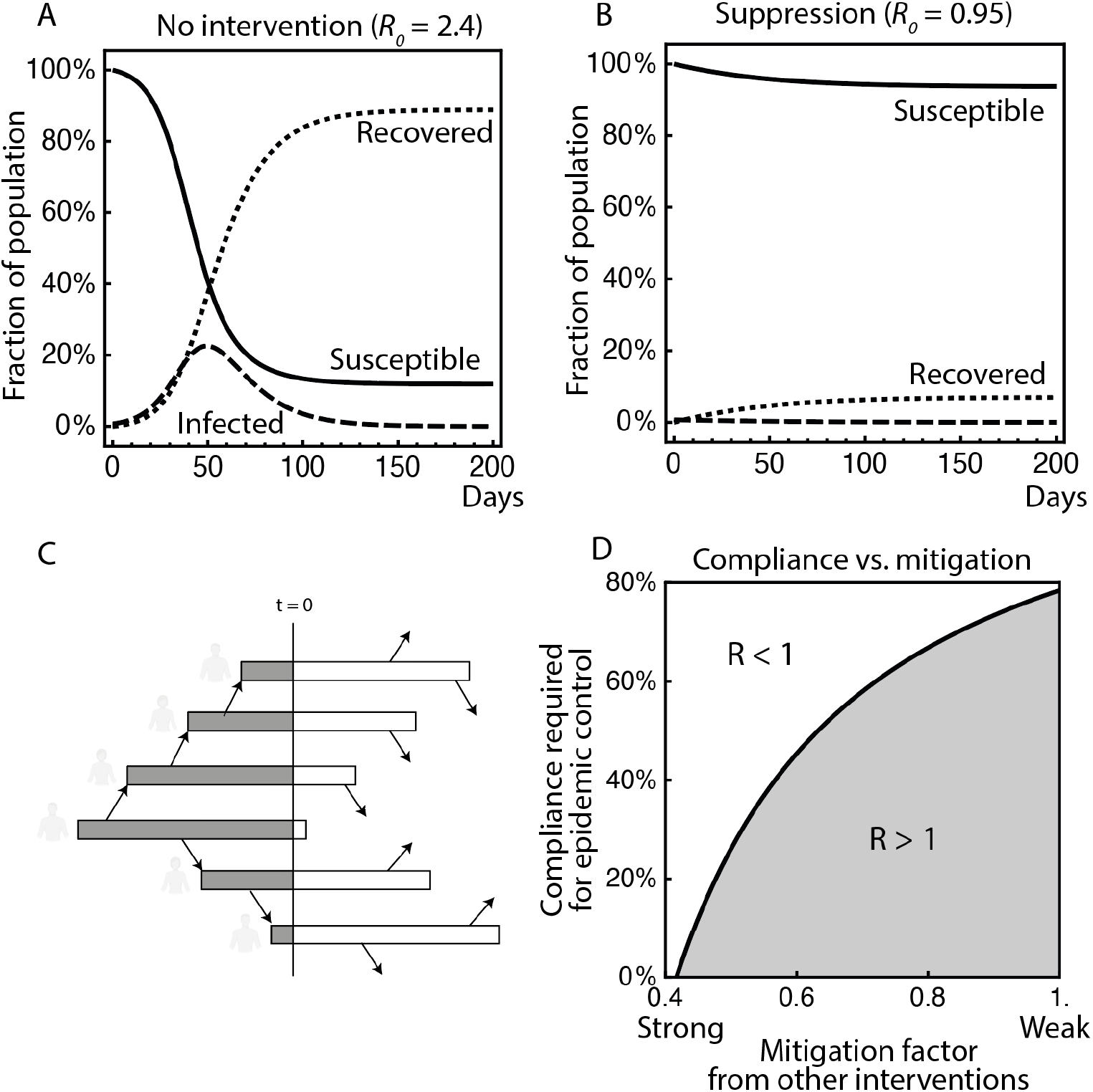
Requirements for epidemic control of COVID-19 using population-scale testing and self-isolation. (A) An SIR model^22^ with *R*_0_ = 2.4 leads to infection of the majority of the population, with a massive peak of active infection that overwhelms the health care system. (B) With strong interventions that reduce *R*_0_ to 0.95, many deaths are avoided. (C) Testing everyone simultaneously cuts all chains of transmission. (D) The required level of compliance (fraction of all individuals) for effective control of the epidemic, as a function of the strength of other interventions and assuming a test with 85% efficacy (fraction of future infections detected). With moderate social distancing, epidemic control can be achieved even with low levels of test compliance.

In the mitigation model, the goal is to reduce R as much as possible but not below 1.0, hoping to end up with a population that is largely immune, without overwhelming the healthcare system in the process (as in **Fig. 1A**, but attempting to flatten the temporary surge of infected individuals). If the virus induces immunity, the mitigation approach also limits the total number of people infected, and leads to “herd immunity” (see, for example Refs.^3,4^), which would limit future epidemics caused by variants of the same virus. However, exponential processes are notoriously difficult to control, particularly in the absence of accurate real-time data and when the effect of policy changes is uncertain. The choice is stark: allowing the disease to spread to a large fraction of a population, however slowly, greatly increases the total number of infected people and would cause a severe loss of life. Furthermore, given the difficulties in controlling exponential processes using limited information, even a strongly enforced mitigation strategy runs the risk of overwhelming the health care system and significantly increasing the mortality rate due to the failure to treat every patient optimally (primarily due to the lack of intensive care capacity). If the healthcare system is overwhelmed, patients must be triaged as in wartime, potentially for extended periods of time.

Notably, both suppression and mitigation are unstable: the mitigation model might first wreck the health-care system and then (as the public demands harsher controls when mortality rises) also wreck the economy. The suppression model might first wreck the economy and then as public pressure forces a relaxation of control, the virus re-emerges. For many months, both approaches are likely to force a large fraction of the population into conditions resembling quarantine. This is because of the large number of asymptomatic carriers of covid-19; in the absence of population-scale testing, the measures need to be implemented in an indiscriminate manner, affecting the whole population. Over time, this will result in severe and unequal economic deprivation. Our estimate is that in the United States, GDP per capita is already lower by about 1000 USD per month, and current estimates for economic loss due to the lockdown range up to $500 billion per month (~ $50 per person per day). Redistribution can offer some protection for the most vulnerable families, but if a loss of income of this magnitude persists for six or twelve months, it could generate a backlash against the social distancing measures that are currently in place.

Given these considerations, we asked if there are less costly and less disruptive alternative strategies to control the pandemic. At low levels of prevalence, testing, contact tracing and quarantine (TTQ) is a very effective means of suppression^3,5^, because it reduces the effective rate of reproduction close to zero. It is not a feasible strategy for suppressing the virus in the current, higher prevalence conditions faced by most countries because it would demand resources that would overwhelm any health department. In addition, the TTQ approach suffers from its own instability. Unless it identifies every single person who becomes infected, asymptomatic individuals that are not identified will generate new clusters that will not be detected until someone develops a severe infection that requires medical care. As a result, once the rate of new cases exceeds the capacity of tracing, even briefly, the epidemic runs out of control and the exponential dynamics make it almost impossible to catch up without imposing a lock-down.

**Here, we propose a radically simpler strategy: test everyone, repeatedly. When someone tests positive, ask them to self-isolate and provide them public assistance that reduces the burden this imposes on them**. This approach relies on a key observation that has not been widely appreciated, namely that what matters is the fraction of all individuals that are identified and isolated. It follows that testing a small number of individuals with a highly accurate test can be much less effective than testing everyone with a less accurate test or reduced compliance. In fact, there is a quantifiable relationship between the reproduction number of a virus, and the efficiency of a population-scale testing strategy that brings the effective reproduction number below 1, as we show below. We use analytical models to derive both an upper and a lower bound on the effectiveness of testing, and demonstrate their real-world relevance using more realistic stochastic models.

The approach has several important advantages. First, it will work no matter how high the prevalence of infection might be. Second, it does not suffer from the inherent instability of contact tracing that requires a very high speed and a secondary mechanism to catch lost contacts. The offsetting disadvantage is that it is a challenge to test at the required scale, but this is not as difficult as it might at first seem. It could be implemented using mass distribution (e.g. regular mail) without returning samples to a central testing site. In fact, the tests required do not even have to be properly “diagnostic.” They only influence the decision to self-isolate. In the worst case, they may cause people who are not infectious to be quarantined, but this is already true for most people in the baseline lockdown scenario. The test can tolerate many false positives, because the result of a provisionally positive test is that someone self-quarantines for two weeks when they did not have to. False negatives are also acceptable as long as people are retested frequently.

The proposed approach is empirical, and does not depend on complex epidemiological models or a highly time-effective, centrally directed response. Its success can be assessed simply by evaluating the test positivity rate over time, and efficacy can be adjusted at an almost arbitrary level of granularity based on positivity rates as a function of other parameters.

To guide initial design, we used the susceptible-infected-recovered (SIR) model to examine the effects of false negatives and noncompliance. We first make the best-case assumption about the timing of the tests: every person who is infected is tested before encountering someone who is susceptible. This limit can be approached, for example, when testing individuals as a condition of release from quarantine, or by a very effective form of contact-tracing. Note that if a perfectly accurate test were applied to the entire population at once, and those who tested positive were fully isolated, the epidemic would immediately collapse with no new infections (**Fig. 1C**). This optimal population-scale testing strategy will succeed if the fraction of infected persons who are isolated exceeds (*R_o_* − 1) / (*R*_0_ − *R_a_*), where *R_a_* is the reproduction number in isolation or quarantine (**Fig. 2A** and **Methods**). For example, if *R*_0_ *=* 2.4 and *R_a_ =* 0.3, and if *p* is the fraction of true positives correctly identified by the test, and *c* is the fraction of the public that complies in the sense that they agree to be tested and follow any instruction to go into isolation, this bound means that the product *cp* must be greater than 2/3.

**Fig. 2.**
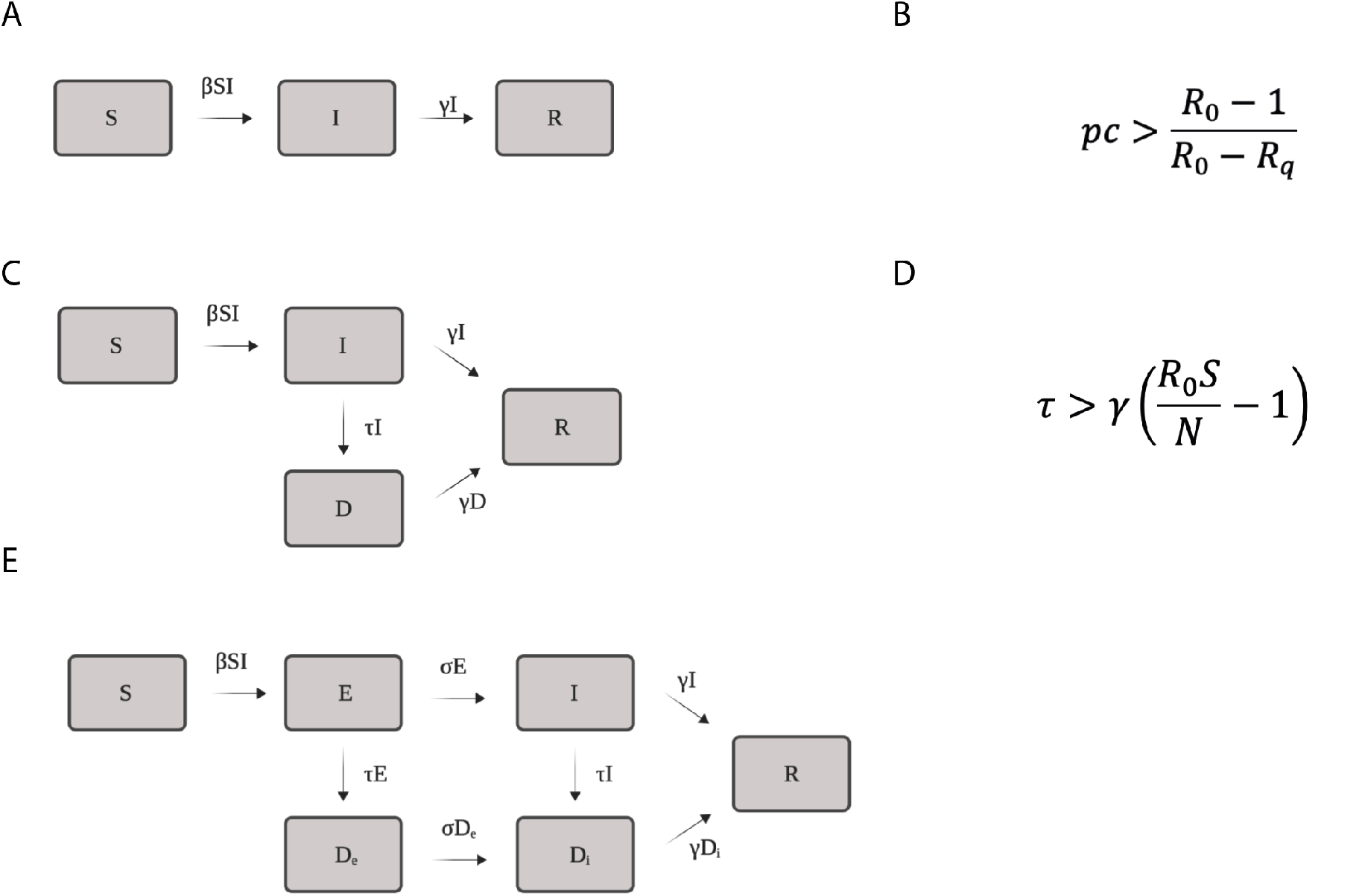
Compartment models and requirements for suppression by testing. (A) Parameters for a standard SIR model. (B) Inequality that must be true to suppress transmission. For the epidemic to collapse, the weighted average of the natural reproduction number *R*_0_ and the reproduction number in self-quarantine *R*_q_ must be less than one. Here, p represents the test true positive rate (fraction of all infectious individuals detected), and *c* the rate of compliance. (C) Parameters for a SIR model with testing and a detected state. (D) Requirements for testing to collapse an epidemic in the SIR model with testing, expressed in terms of the testing rate *t* required in a population where all individuals are susceptible, with inverse infectious interval λ. (E) Parameters for the discrete, stochastic SEIR model.

Next, we assume instead that the test sensitivity and compliance are perfect, so *p* = *c =* 1, and consider the worst-case assumption on the timing of the tests: each day, a randomly selected fraction of the population is tested. Under that strategy, we find that testing at a rate greater than a fraction (*R*_0_*S/N* − 1) of the population per infectious period will ensure that *R*_e_ < 1 (**Fig. 2B**, **Methods**; S/N is the fraction still susceptible). Real-world testing strategies could do much better than test at random, for example by implementing procedures that test individuals concurrently within a region; that run the screen as a sweep across a country; that slice the population into groups that are tested in a cycle; that test individuals that have many contacts, or use other variables to predict who is more likely to be infected or more likely to infect others and to test them more frequently (see **Methods**). The theoretical limit of performance of most testing strategies lies between the two bounds delineated above. The order of performance is defined by a simple order rule (see **Methods**), and is: testing the vector (e.g. blood before transfusion), testing before infectiousness, testing all people at the same time, testing each person but at different times, and random sampling of individuals. Cyclic processes that occur naturally (e.g. activity cycle across a week) or due to policy (e.g. rolling lockdown; Ref. 29) will also partially synchronize infectious intervals, and facilitate the application of the stronger testing regimes. Furthermore, if local or population-scale information exists that can be applied for contact-tracing, the efficacy of the regime can be significantly improved by isolating or quarantining the contacts of the individuals who test positive.

The fact that the approach will work is clear if one considers that the current approaches are “natural” variants of the population-scale testing regime. For example, lockdown corresponds to a test with a sensitivity of zero and false positive rate of one, and isolation of symptomatic individuals corresponds to a a non-biochemical test that measures presence of covid-19 based on self-assessment of generic symptoms, leading to a relatively low sensitivity and specificity, and a suboptimal timing (self-assessed or even expert-guided symptom-test for covid-19 is centered at the middle of the infectious interval, and will not detect cases that remain non-symptomatic throughout the infectious period). These parameters can clearly be improved by an introduction of a population-scale biochemical test. The rates of testing that are required to suppress an epidemic are sensitive to the input parameters. However, despite the exponential nature of the epidemic itself, the sensitivity of the required testing interval to variation in most parameters is linear or nearly so. For example, both a shorter infectious period and a larger *R*_0_ shorten the require testing interval proportionately. The most impactful parameter is the delay in obtaining a test result *d*, which should be minimized in any realistic plan for population-scale testing. This is because if *d* is longer than the sum of the detection lead time (*l*) and *R*_0_ divided by the infectious period, the epidemic cannot be collapsed by any rate of testing in the absence of other interventions (see **Methods**).

Population-scale testing positively interacts with other strategies. Interventions that reduce *R*_0_ — e.g. working from home, improved hand hygiene, isolation even with mild symtoms, the use of masks and social distancing — are additive with respect to the testing, and hence lower the required frequency of the tests. Similarly, as the epidemic progresses and fewer people remain susceptible, the frequency of testing required to control the epidemic drops. The relationship between these variables in the SIR model with testing is captured by the following inequality (**Methods**), which relates the testing rate (τ, average fraction of population tested per day), the recovery rate (γ, inverse of the infectious period), and the proportion still susceptible (S/N):

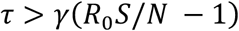

As shown in **Fig. 3**, with γ = 1/5, assuming moderate non-pharmaceutical interventions that reduce *R*_0_ to the range 1.2 – 1.5, and assuming most (90%) of the population is still susceptible, population-scale testing on average every 15 – 120 days would be sufficient to bring the effective reproduction number below 1.0 and thus control the epidemic. With much longer (14 days) or shorter (3 days) infectious interval, the required rate of testing changes accordingly. Importantly, in a realistic range of parameters, it is particularly difficult to find values that would both make it impossible to suppress the epidemic by population-scale testing, and prevent relatively easily implementable alternative means of suppression (wherein testing would commonly also be an important component). For example, if the infectious period was only two days, suppression of the epidemic by random testing alone would be difficult – but instead a short quarantine followed by a test would be highly effective. Similarly, if individuals with short and long infectious intervals co-exist in the population due to genetic or environmental variation, the short duration infections can be suppressed by lockdown, and the long ones by testing. The figure also illustrates the trade-offs involved: if population-scale testing rates can be increased from once every four months (1/120^th^ of the population per day) to once per two weeks (1/14^th^ of the population per day), then alternative measures such as lockdowns can be reduced from *R*_0_ ≈ 1.2 to *R*_0_ ≈ 1.5.

**Fig. 3.**
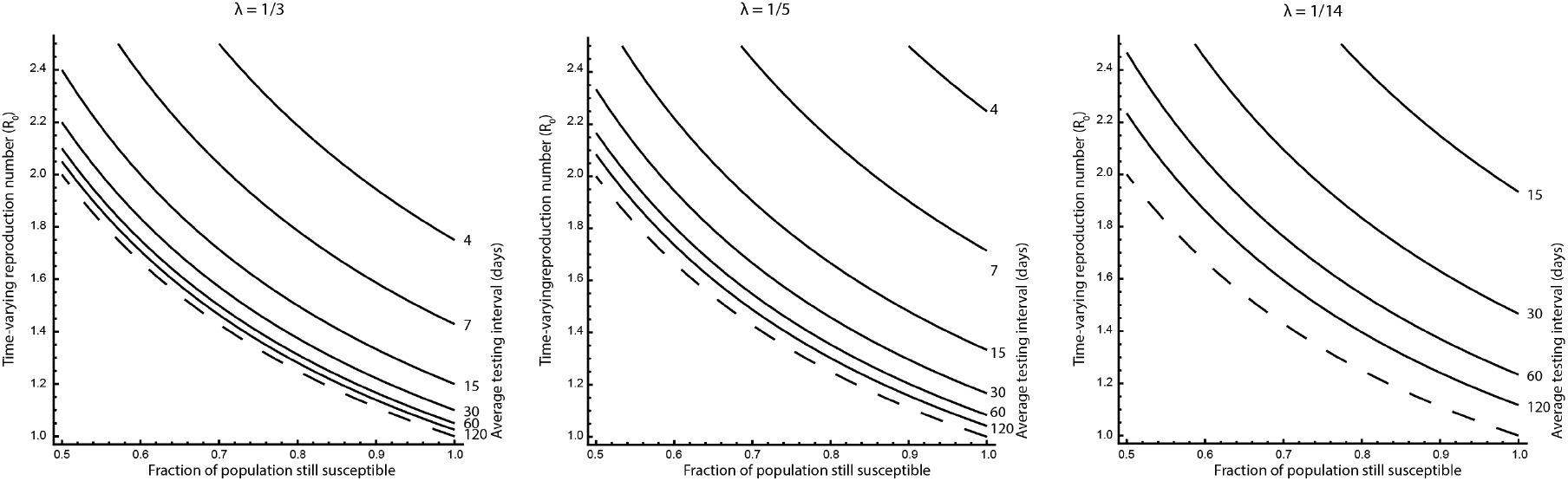
Testing frequency required to control transmission as a function of the fraction still suceptible (*S/N*, horizontal axes) and the reproduction number (*R*_0_, vertical axes) for three recovery rates corresponding to 3, 5 and 14 days infectious periods. Solid curves, testing frequencies, indicated as average number of days between tests, per person. Dashed curves, herd immunity threshold below which transmission collapses without testing.

The standard but simple and deterministic Susceptible-Infectious-Recovered (SIR) models used to calculate these bounds is based on strong assumptions and approximations, such as random mixing of all individuals, perfectly accurate testing, full compliance and perfect isolation of cases. To relax those assumptions, we implemented a more realistic numerical simulation using a stochastic model with testing and an exposed state (i.e. a stochastic SEIR model). The deterministic model above predicted that testing at least every ten days would be required to control an epidemic when *R*_0_ *=* 1.5. **Fig. 4** shows a simulation that starts with 100 infected individuals out of a 100,000 population, with the infection fatality rate was set to 1%. Population-scale testing using random tests at a rate of once per ten days immediately suppressed transmission as expected, with 0.03% of the population dead. In contrast, without testing, viral spread caused a surge of cases that eventually led to herd immunity with 0.58% of the population dead. Translated to a US-sized population, more than 1.5 million lives were saved.

**Fig 4.**
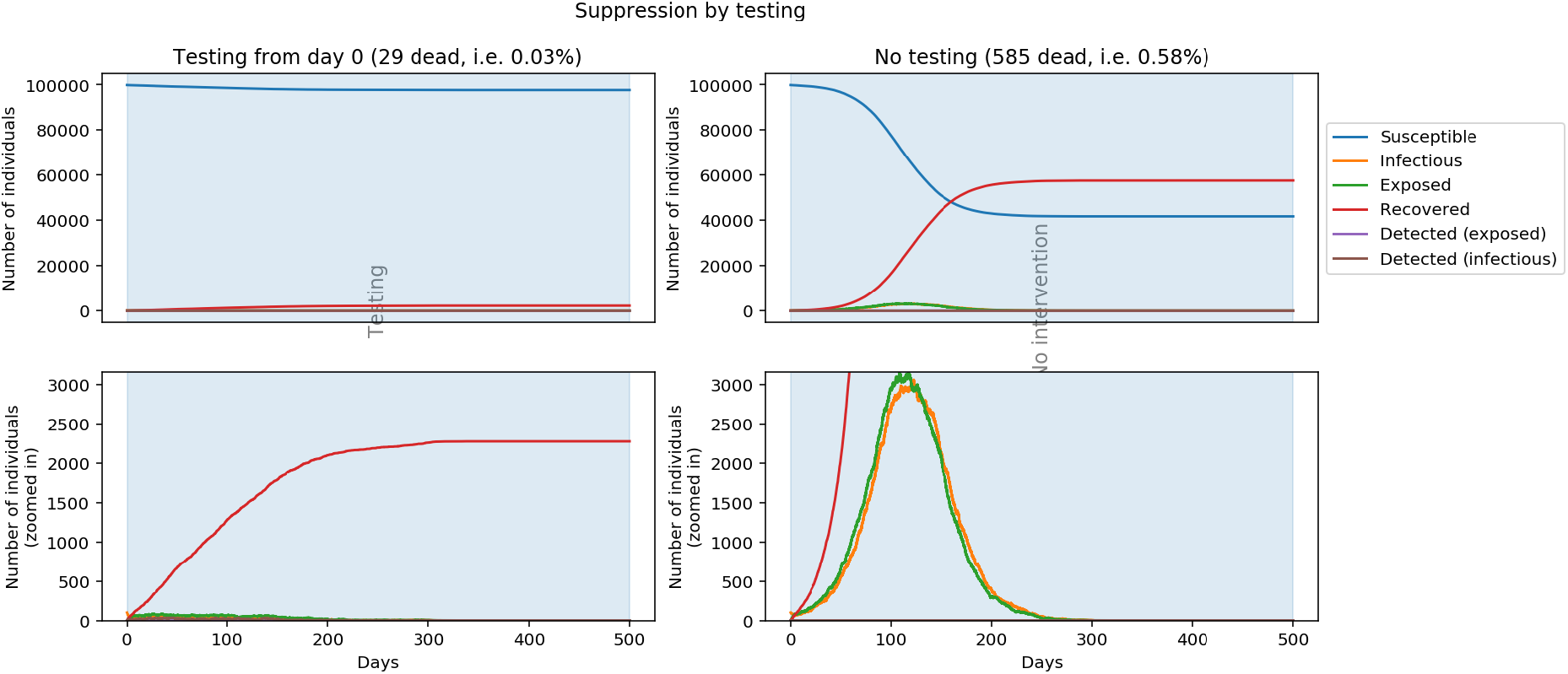
Simulation of suppression by testing using the stochastic SEIR model. Simulations with (left) and without (right) testing at a rate of one test per ten days (τ = 1/10) were started with 100 infected individuals, using *= 0.2, σ =* 1/5, *γ =* 1/5 correspodning to five days infectious period and *R*_0_ *=* 1.5. Testing was assumed perfect and test-positive individuals were perfectly isolated. The infection fatality rate was 1%. The total population was set to 100,000 individuals. Note that without testing, the epidemic reaches herd immunity with fatalities that scale with population size, whereas with testing, the epidemic is extinguished and the number of fatalities scales with the size of the initial outbreak regardless of population size.

Equally importantly to considering the probability of success, a decision to apply a treatment or implement a policy must weigh the consequences failure. In the general case where testing is insufficient and *R*_0_ remains above 1 despite all interventions, the benefit from the testing would be the difference between the attack rates at original *R*_0_ and *R_eff_* under the testing regime (**Fig. 5**). As is clear from the figure, even when testing is insufficient to fully suppress the epidemic, testing at any rate reduces the final number of infected and deceased individuals. We conclude that the consequence of failure to fully implement the policy would still lead to improvement in both public health and economy, a finding that does not depend on the fractional scale at which the policy is implemented. We therefore see no medically or economically justifiable basis for not increasing Covid-19 testing to the maximum scale permitted by any limiting technical, logistical or economic constraint.

**Fig 5.**
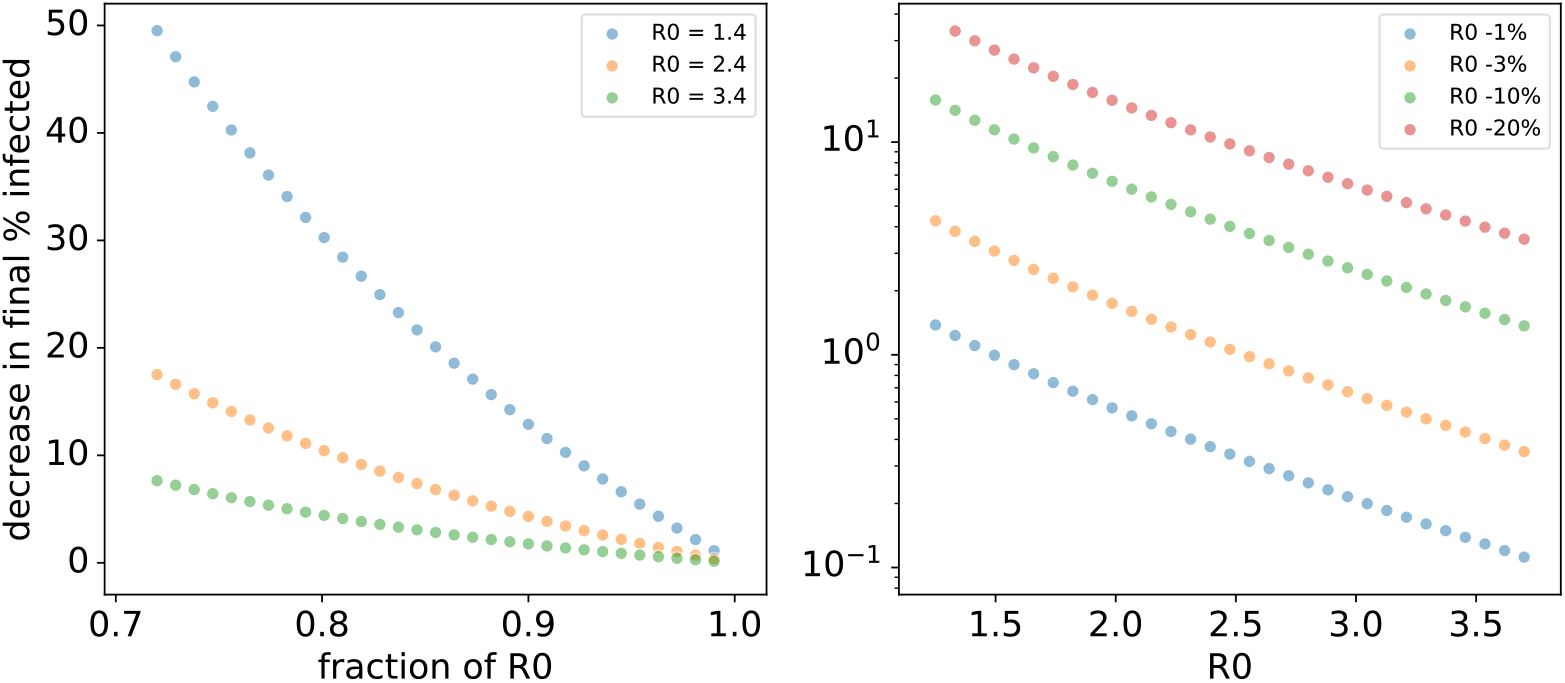
Consequence of failure to suppress the epidemic by population-scale testing. Left: Decrease in the final attack rate (y-axis) as a function of change in *R*_0_ induced by population-scale testing (x-axis). Note that the effect is nearly linear, and that effects of up to 50% of the total population can be observed even when *R*_0_ remains over 1. Right: Decrease in the final attack rate (y-axis) as a function of initial *R*_0_ that is decreased by 1 to 20% by population-scale testing. The final attack-rates (z) were calculated using the lambert W function: z = 1 + *W(-R*_0_ exp(-«_0_))/K_0_.

There are many plausible technical approaches to population-scale testing. Such a test can be based on presence of virus antigen, either in the form of viral proteins (technically more difficult but possible^10^), or viral RNA like current state-of-the-art diagnostic tests (for example Ref. ^11^). In both cases, a home self-test is preferable due to the simple logistics and quick time to result, which reduces the crucial latency to isolation of infectious individuals. Despite the technical simplicity, it is difficult to translate current tests designed for medical diagnostic purposes to a field setting. Current diagnostic tests for SARS-CoV-2 are qRT-PCR assays that require (1) nasopharyngeal swab collected by a trained nurse, (2) sample collection in viral transport media, (3) RNA purification, (4) reverse transcription and quantitative PCR. The test is highly accurate, and the total cost is in the order of $100. Such highly accurate testing is critical for accurate diagnosis of infections in a hospital setting. However, due to the very detailed and specific regulation, specialized staff and equipment, and centralized testing facilities, such tests have proven difficult to rapidly scale above thousands of assays in each location. A distributed system of sample collection and testing could, however, conceivably be used to scale qRT-PCR to population levels, particularly when using a regional sweeping approach to limit the number of simultaneous tests needed. The capacity could also be increased 10 to 100-fold by group testing^24^, a method with a long history of use in public health that was originally designed for Syphilis tests, and now commonly also used for optimally efficient detection of defective components in industrial production.

A parallel relatively centralized testing method based on existing DNA sequencing technology could also be fielded rapidly. In this approach, viral RNA in the samples is used to generate DNA sequences containing the virus sequences, a sample DNA barcode (to identify each case) and two unique molecular identifiers^23^ at both ends of the resulting DNA fragment (to count the number of virus RNAs per sample and to ensure that patient samples do not get mixed in the reaction), and then sequenced using a massively parallel sequencer. This approach is very scalable, as in principle, a single sequencing instrument that is routinely used in scientific research can report more than a billion results per day. Furthermore, in the future, a test based on sequencing^19-21^ that covers many acute infections could also be used to suppress or even eradicate a large number of infectious diseases simultaneously. This would be very difficult to achieve using vaccines or drugs that target each infectious agent separately.

Alternatively, we envisage supplementing the current testing regime with a mass-produced home test kit that could be used by anyone, result in a simple easily-understood readout, and be performed without specialized equipment. The test should be as easy to use as a pregnancy test, to ensure maximal compliance; importantly, using a home test, the time delay to report the result *d* would be effectively eliminated. Boxes of e.g. 50 tests would be mass-mailed to all citizens, and a national information campaign would encourage everyone to test themselves frequently. In an infected individual, viral RNA is present at reasonably high levels in nasopharyngeal swabs, throat swabs, sputum, and stool for up to two weeks^12^, with the greatest amounts in saliva, sputum, stool. Saliva might be the ideal source for a home test kit, given the ease of sampling.

Tests suitable for home use are already in development. For example, an isothermal and colorimetric test has been described^13,14^, based on reverse transcription-loop mediated amplification (RT-LAMP) technology. This test has several desirable properties: unlike PCR, it does not require temperature cycling; the readout is binary and can be achieved by simple observation; and it can start from crude samples^15^. Many other technologies also have the potential to detect viral RNA rapidly and isothermally^16-18^; these include recombinase polymerase amplification (RPA), transcription mediated amplification, nicking enzyme amplification reaction (NEAR), rolling circle replication, and *in vitro* viral replication assays. Finally, lateral-flow strips for the detection of viral antigen have been announced, although their performance has not yet been assessed.

We have outlined a framework for population-scale testing as an effective intervention, and derived formulas that relate the key design parameters for such a strategy. Importantly, the benefits of such a strategy are likely to greatly outweigh the cost, even in the event that it would fail to fully suppress the epidemic.

## Data Availability

The paper is computational and therefore includes no new experimental data

## Acknowledgments

We thank many colleagues for comments on the early version of the work. We are especially grateful to Drs. Minna Taipale, Mikko Taipale and Paul Pharoah for review of the draft of manuscript. A draft of this paper was initially released as a public preprint (Ref. 25), and supporting, independently developed model was reported by P.R. on www.paulromer.net. We also note that during the writing of this work, we became aware of two independent analyses, one by Julian Peto (Ref. 26), and the other by a team consisting of David Berger, Kyle Hirkenhoff and Simon Mongey that report similar conclusions (Ref. 27).

## METHODS

### Assumptions, parameters and choice of models

The present work relies on two principal assumptions: 1) testing an individual for the presence of viral RNA or antigen will predict future infections originating from the same individual, and 2) future infections can be prevented by isolating the individual. If these two assumptions hold, an epidemic will collapse if the testing regime can decrease the rate of generation of new infections below the rate of recovery or death of the infected individuals. If the epidemic does not collapse, the testing regime will still decrease the rate of generation of new infections, leading to a less dramatic but still beneficial outcome. It is important to note that the above conclusions rely only on the two assumptions stated above, and not on the specific model used to assess the quantitative relationships between the testing rate and design, and summary statistic abstractions such as *R*_0_.

The epidemic was first modelled with a standard (continuous, deterministic) susceptible, infected, removed (SIR) model. An important consideration in the choice of a model to represent underlying physical reality is that it represents the level of abstraction that is relevant to the scientific question addressed. The SIR model was chosen as it is sufficiently abstract to capture general features that operate at a population-level, and does not fit to particular conditions in specific countries. Furthermore, the abstract nature of the model is further justified by the uncertainty in the input parameters, inclusion of each of which would require a new set of separate assumptions. Just as an example, we lack accurate parameters that describe the rate of test positivity as a function of future infectivity of an individual. Using any individual parameter that includes uncertainty introduces error, and using any pair of separately measured parameters that depend on each other introduces further error. Therefore, the modeling we perform here is solely aimed at evaluating the general feasibility of the approach, and for setting initial parameters that should be dynamically adjusted during the intervention itself.

It is important to note that parameter ranges where testing cannot effectively be used to suppress an epidemic can be found, particularly when combining worst-case estimates across studies. The weakest performance of testing is in cases with extremely high *R*_0_, or very long incubation period with low viral load, followed by rapid increase and high infectiousness (however, long incubation period is not likely due to the initial exponential replication of viruses in the host). The failure of some countries to suppress Covid-19 by the means available to them also suggests that some initial parameter estimates are unlikely to be correct (e.g. initially it was estimated that infectious period starts after symptoms). This also indicates that a different approach is required than what was initially taken, and that asymptomatic cases need to be identified to control spread.

One important parameter that affects performance of testing is the false negative rate of tests, which varies mainly due to viral load and sampling errors. We would like to note that methods based on amplification of nucleic acids generally have higher sensitivity to detect (a correlate of) an infectious viral particle than the viral infection itself. This is due to three reasons: First, viral infection of cells is a complex process, that tends to be inefficient, with many viruses needed to establish one productive infection. Second, volume of a sample taken from a patient (microliters) is generally much higher than the total volume in the particles that reach the airways of the infected patients (nanoliters or picoliters; Ref. 30). Third, nucleic acid detection methods can also detect RNA released from dead cells, RNA present in dead viruses, and in the subset of defective interfering viral particles that carry the relevant segment of RNA. Despite these advantages, even PCR tests return false negatives for clinically diagnosed cases. However, the false negative rate estimated in the literature (e.g. 71% according to Ref. 31) cannot be easily used in analysis of the effectiveness of population-scale testing. This is because the rate of false negatives is estimated from all cases, and not from infectious cases. Using two independent measured values for infectiousness and false negative rate leads to measurement of a form of lower bound for the relevant variable (time-dependent probability of obtaining a positive test result as a function of the area under the curve of future infectiousness).

### SIR model

In addition to the very general assumption that there are a relatively large number of cases, which allows modeling of a partially discrete system using a continuous model, the SIR model is based on the following standard assumptions: (1) the population is fixed, (2) it mixes homogenously, (3) the only way a person can leave the susceptible group is to become infected, (4) the only way a person can leave the infected group is to recover from the disease, (5) recovered persons become immune, (6) age, sex, social status, genetics etc. do not affect the probability of being infected, (7) there is no inherited immunity, and (8) the other mitigation strategies and testing are independent of each other (for **Fig. 1D**). The assumption (2) leads the SIR model to overestimate viral spread, as in reality population has substructure (e.g. families, workplaces) and is geographically separated and contacts are more likely between subsets of the population; this is not expected to materially affect our analysis as our conclusions are not based on the absolute rate of the spread, only on its exponential nature. Furthermore, the SIR model is extremely conservative in the sense that it overpredicts the rate of growth of the infected population when the number of infected individuals is low; it does not even allow for extinction of the virus – a very desirable outcome, which obviously has a non-zero probability of occurrence in any real-world scenario.

In addition, we modeled the effect of testing in two ways. The first, maximally effective testing strategy assumed that every individual was tested before they infected another person, leading to the upper bound on testing performance in **Fig. 2A-B**. Under this model, the requirement for collapsing the epidemic is that the weighted average of the basic reproduction number *R*_0_ and the reproduction number in isolation or quarantine *R_a_* must be less than one:

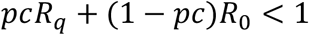

i.e.

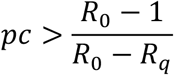

Here, *p* is the true positive rate of the test and *C* is the compliance (fraction of all tested individuals who actually self-isolate).

Using *R*_0_ *=* 2.4 and *R_q_ =* 0.3 for COVID-19, the product of the true positive rate and compliance must be greater than two thirds:

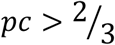

The second, lower bound testing strategy (**Fig. 2C-D**) was modelled by adding an additional ‘detected’ state to the model, and adding transitions from infected to detected (with rate *τI*) and from detected to recovered (with rate *γD*). This corresponds to continuous random testing of the population at a fixed rate *τ* per person per day. Here, the requirement for successful collapse of the epidemic is given by the basic reproduction number (assuming perfect isolation; **Fig. 2D**), as follows. First, the rate equations for the SIR model with testing are:

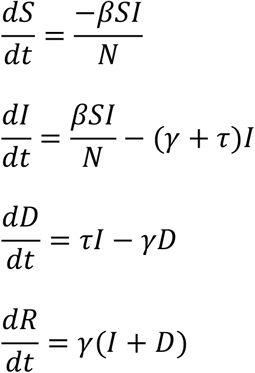

Rewriting the second equation above as follows:

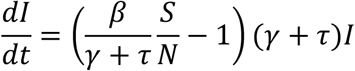

makes it clear that *dI/dt* will be negative (i.e. the epidemic will collapse) only if:

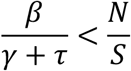

Note that the ratio *β/γ* is the basic reproduction number *R*_0_, so that the previous inequality can be rewritten as follows:

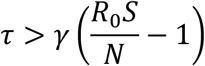

In other words, the testing rate must exceed a threshold given by the recovery rate *γ* (inverse of the infectious period), the time-varying reproduction number *R*_0_ and the fraction of susceptible individuals *S/N*. The required testing rate drops if non-pharmaceutical interventions reduce *R*_0_. Similarly, as the epidemic progresses, the required testing rate drops as fewer and fewer individuals remain susceptible and herd immunity kicks in.

### Stochastic SEIR model

The SIR model fails to account for several key properties of real epidemics, such as social and geographical population structure, the discrete and stochastic nature of infection and disease progression, and the fact that testing cannot be instantaneous. To account for such more complex real-world phenomena, we implemented a stochastic network model using the Gillespie algorithm for accurate numerical simulation of the stochastic dynamics. We used the seirsplus Python package (https://github.com/ryansmcgee/seirsplus), which models a population where each individual transitions between six states: susceptible, exposed, detected-exposed, infectious, detected-infectious, and recovered. The two detected states are used to model the effectiveness of testing and isolation. The population consisted of 10,000 individuals. Detailed source code with comments and parameter settings for each model are available in the accompanying Jupyter notebook at https://github.com/paulromer149/ubiquitous-testing.

### Efficiency of testing

To understand the efficiency of testing, it is helpful to consider three separate processes: the process of the infection itself, the ordering of the tests relative to the infectious process, and the structuring of the testing process.

The infection itself can be divided to four periods: the period that an individual carries a virus to four consecutive intervals: undetectable (u), lead time for testing (l), infectious period (i) and antigen positive but non-infectious period (a). In a model of testing, the classical incubation period has to be split into two (u+l), because the relevant interval for antigen/RNA testing to suppress an epidemic by isolation of the tested individuals is not the infectious period i, but l+i; l exists because of the very large difference in volume transfer between a test and an infection in the case of a respiratory virus. The period relevant for contact-tracing is even longer, l+i+a, as detection of the recent infection leads to identification of a starting point for the search of currently infected contacts. It is important also to note that the intervals are abstractions of infectious and non-infectious virus genetic material / particle concentration over time within an individual; this is never 0 in an individual who carries the virus, and varies in both time and magnitude, affected by the precise dynamics of the infection process itself.

To understand the difference between testing (1) the vector, (2) prior to transmission, (3) everyone at the same time, (4) everyone in a time separated manner, or (5) the population by random sampling, it is helpful to consider extreme case of certainly and completely collapsing an epidemic by testing and isolation, using a perfect test that detects all infected individuals, and complete isolation. For optimally achieving this, it is necessary to identify everyone who is infected before they have infected anyone else (denoted efficiency, e = 1) and quarantining every infected individual (c = 1). This requires obtaining a minimum of n bits of information for a population of size n. Note, however, that a single test performed as a group test can return more than 1 bit, and that this can be useful in detection of super-spreaders with very high viral titer (Ref. 32). The order of performance of different strategies can be derived from the following simple model:

Let there be a constant number of tests (**T**) per individual placed on sequences consisting of four elements: I_0_ (incoming infection of individual 0), **T** (test), I_n_ (infection of individual n, with I without index indicating any infection) and R (recovery of the individual).

All sequences in case (1) have the following form:

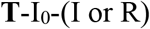

This allows the test to prevent even the first infection, which is clearly the best possible way to place a single test per individual.

All sequences in case (2) have the following form:

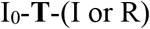

This cannot prevent the first infection, but is clearly the second-best possible way to place a single test per individual.

Case (3) allows the second-best order:

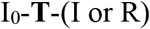

but also recursively:

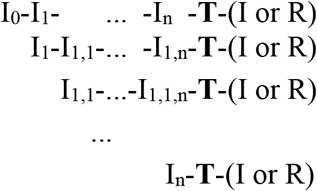

note that the concurrency ensures that tests are placed on each interval optimally (one on each branch of the tree).

Whereas case (4) can allow the above sequences, and in the absence of specific ordering of infections and tests:

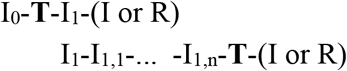

which places two tests along the same chain, and also recursively:

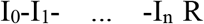

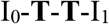

A branch with no tests at all. Ordering of tests and infections, by for example sweeping a region and preventing movement of individuals in the opposite direction improves this strategy, but its performance will remain below case (3) because the branches will grow until a test is placed on them.

Case (5) performs the worst, because it also allows:

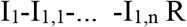

Which tests the same individual twice during one infectious period. This is in itself good, but decreases overall efficacy of the regime as it leads to more branches with no tests than case (4).

Case (3) is the weakest testing regime that always achieves complete collapse; this is achieved by testing everyone at the same time with a perfectly accurate test that returns one bit (positive or negative). In this case, e = 1 and c = 1. However, when tests are separated in time (Case 4), the order of testing becomes important. Most strategies for testing n individuals during time t_test_interval_ before t_0_ have e < 1, and are not sufficient to completely collapse the epidemic using one testing round, as e depends on the relationship between the order of testing and the order of infections. For example, using a random order of testing allows some individuals that have already been tested negative to become infected during the t_test_interval_ (the mutual information between test results and person being infected at t_0_ is less than one bit). However, some other regimens using a perfectly sensitive test can collapse the epidemic (but not always prevent all future infections): for example, a geographical sweep where infections (individuals) are prevented from crossing a moving test front can be used to identify every infected individual in the population by performing a single round of n tests.

In case (5), random sampling of n individuals, e is always less than 1. The testing becomes less efficient than testing each of the n individuals at the same time, because some individuals are tested twice, and some not at all; some information is thus not obtained, and some tests do not return information that is completely independent of information returned by other tests (sum of mutual information between all pairs of tests is not 0 bits). In other words, if individuals are selected randomly, during a given time interval, the tests will miss some individuals, and some individuals are tested more than once (this increases true positive rate for those individuals, but this does not make up for failing to catch some individuals entirely).

Considering the extreme case of immediate collapse, it may appear that testing in a time separated manner or by using random sampling will not work because non-concurrent testing can permit infections to cross the testing boundary, and random sampling clearly leaves some cases undetected. However, this very intuitive idea is incorrect, as collapsing an epidemic only requires that the rate of generation of new cases per current case is less than one. The limit for random testing can be obtained using the SIR model extended with testing (SIR+T), which abstracts away individuals and thus can (only) be used to investigate the effect of random, time-separated testing. Analytically from this model, as shown above, the *R* < 1 condition is true when tests are performed at a rate that is higher than *R*_0_ − 1 tests per mean infectious period. The same limit results from the consideration of a model that is similar to the the simple model above: reducing *R*_0_ to less than one using the method representing the lower bound – a completely random testing regime – requires that an infected individual has less than an equal probability of (a) infecting another individual over (b) being tested and isolated or recovering from the infection (analogously, in SIR+T, the combined testing and recovery rate needs to be higher than the rate of new infections). Events (a) can recur, but either event (b) terminates the chain. Therefore, at R = 1 there will be on average one (a) event, which requires that the order of the infectious and protective events are randomly ordered with respect to each other, with equal density. This yields *R*_0_ — 1 tests and one recovery per *R*_0_ infections per infectious period, and an upper limit of *R*_0_ tests per infectious period at infinity (because as *R*_0_ → *∞* the expectation value for the number of test required per *R*_0_ becomes the geometric series 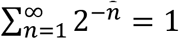).

Outside of the theoretical consideration of *e* = 1, multiple population-scale tests are always required to collapse the epidemic in the absence of other interventions that achieve the same aim. Performing multiple tests over time imposes an additional constraint on optimality – the allocation of tests to each transmission interval. As described above, best performance of continuous testing and isolation is thus achieved when testing is performed immediately after infection for each individual, or as requirement for exiting quarantine. Testing at border crossings, conditional opening of lockdown, or some regimes that apply contact-tracing may come close to approximating this limit, which for Covid-19 is *pc* = (*R*_0_ − 1) / *R*_0_ *=* 0.57 per mean infectious period; **Fig. 2**). However, in most scenarios, such testing efficacy is difficult to maintain over time (because contact is lost, and the unknown infectious intervals rapidly become randomly distributed over time). This level can thus be considered an upper limit of performance of any scenario applied at population scale.

Using a test whose true positive rate is 1 and testing everyone at the same time performs as well as the optimal strategy. As test sensitivity decreases, the performance of the concurrent regime becomes lower than optimal. However, concurrent testing still performs well above the lower limit obtained from the random testing model. The required pc rate to bring *R*_0_ < 1 using concurrent tests has a simple relationship with the exponential growth of infectious cases. Over interval t-t_0_, pc > 1-(infectious cases at t_0_)/(infectious cases at t). However, it is not as simple to relate this to original *R*_0_, because the relationship between *R*_0_ and growth rate is a function of the distribution of the generation intervals (Ref. 28). Estimating at *R*_0_ = 2.4 using even probability distribution of infections over time, the infected population becomes approx. eight times larger in a single infectious period (for SIR, this would be different due to the different assumption about the distribution of infectious periods; see Ref. 28). This means that a testing regime that is regularly spaced at 14 day intervals should have pc value of > 7/8 = 0.875 to bring *R*_0_ < 1. This is confirmed using empirical simulations to assess the rate of exponential growth in the complete absence of immunity and all other types of interventions; the limit *R* = 1 at *R*_0_ = 2.35 with testing every infectious period is reached when *pc* ≈ 0.85 (compared to 0.58 for testing each individual directly after infection). The required testing interval at *R*_0_ *= 2.35* and *pc =* 0.8 in the absence of other interventions and immunity is ~ 0.8, 0.6 and 0.4 times the infectious period for concurrent testing, testing each individual randomly once during each testing period, and continuous random testing, respectively.

These considerations can be summarized as follows: the order of testing efficacies is: all vectors (at edges of network) > everyone before they have had a chance to infect anyone > everyone at the same time > everyone once during a period > testing by random sampling – with population-scale testing remaining feasible and cost-effective by one or more orders of magnitude across all these regimens.

For the structuring of the testing process, one should carefully consider the entire testing regime, which is invariably population-scale for all infectious diseases, even if this intuitively may appear not to be the case. In practice, many different approaches are used, but most behave formally as consecutive testing, with the increased number of layers commonly (optimally in any real situation) increasing both the true positive and the false negative rate. This is because the first test (e.g. appearance of symptoms, or being a contact or not) can increase the true positive rate, but also determines the lower bound for the false negative rate of the whole process (e.g. in a purely contact-tracing based approach, infection of a missed contact cannot be detected biochemically). Thus, it is important to understand that while predictive tools such as appearance of symptoms or contact-tracing can increase the true positive rate, this invariably comes at some cost of increase in the false negative rate (e.g. asymptomatic individuals, lost contacts). Many predictive approaches are also highly time-dependent; for example, timing of tests in contact-tracing optimally approaches the best case (testing before infectivity), but in the worst case performs far worse than random testing (testing after an individual has already infected everyone they can). Thus, due to the increase in false negative rate, a contact-tracing approach must both be combined with another mechanism to catch the escaped contacts, and performed extremely fast to improve the timing of the tests. In summary, and in simple terms, any consecutive approach always – by definition – will have a time-cost that a population-scale biochemical testing approach will eliminate.

